# Integrative single-cell analysis of epigenomic and transcriptomic states in patients with systemic sclerosis-associated interstitial lung disease

**DOI:** 10.1101/2023.09.05.23294979

**Authors:** Shifang Li, Meijiao Gong

**Affiliations:** Laboratory of Immunology and Vaccinology, FARAH, ULiège, Liège 4000, Belgium. SF and MJ share senior authorship

## Abstract

Interstitial lung disease is the primary cause of death in individuals who have systemic sclerosis, one of the autoimmune connective tissue diseases. Understanding the pathophysiology of the disease is crucial to developing treatment options. Here, we performed a single-cell multi-omic analysis on lung tissue samples from patients with systemic sclerosis-associated interstitial lung disease (SSC-ILD), profiling chromatin accessibility and gene expression in the same samples and discovering significant cellular heterogeneity. Systemic-venous endothelial cells (ECs) have been shown to be pro-inflammatory and highly active. In addition, it was shown that the transcription factor FOSL2 targets the genes involved in response to unfolded proteins in systemic-venous ECs. Furthermore, we prioritized functional risk variants for systemic sclerosis using a genome-wide association study. Ligand-receptor analysis revealed that ECs significantly increased interaction with B cells via CXCL10-CXCR3 in patients with SSC-ILD. Overall, our analysis emphasizes epigenetic and transcriptional patterns in systemic-venous ECs, which might be beneficial in understanding the pathogenesis of SSC-ILD.

## Introduction

Systemic sclerosis (SSC) is a multi-organ disease with clinical symptoms such as skin and internal organ fibrosis and vascular abnormalities. Because of limited therapy choices, SSC causes significant morbidity and mortality. One of the primary causes of SSC-related death is interstitial lung disease (ILD) [1]. However, the processes underlying fibrosis in SSC-associated interstitial lung disease (SSC-ILD) remain largely unknown. According to research, epithelial cell damage, activation of innate and adaptive immunity, and recruitment and activation of fibroblasts may all contribute to the excessive synthesis of extracellular matrix and scar formation in SSC-ILD, similar to the mechanisms in other fibrotic lung diseases [2]. Furthermore, in SSC-ILD, myofibroblasts have been demonstrated to be essential effector cells for extracellular matrix remodeling [2–3]. Recent research has given more insight into the role of SPP1 macrophages in pro-fibrosis in SSC-ILD [4]. However, the role of the diversity of other cellular subpopulations found in healthy and SSC-ILD lungs in shaping SSC-ILD pathophysiology is unclear.

Single-cell sequencing, which includes single-cell RNA sequencing (scRNA-seq) and the assay for transposase-accessible chromatin sequencing (scATAC-seq), is a valuable method for investigating the molecular and cellular heterogeneity of cell types. EC, one of the innermost cell types lined along vessels in all organs and tissues through hosts, is also in the bloodstream, where it can modulate inflammation by regulating immune cell transport, activation status, and function [5–7]. Furthermore, EC in the lungs and liver not only promotes immune homeostasis but also mediates the balance between tolerance and inflammation [6]. However, the role of endothelial cells (ECs) in lung fibrosis is not well-defined. A previous study showed that ECs to mesenchymal phenotypic change was observed in the lung tissues of SSC-ILD patients [8]. A recent study found that ECs in the lungs regulate pulmonary fibrosis via FOXF1/R-Ras signaling [9]. However, our understanding of EC heterogeneity in SSC-ILD at the single-cell level remains limited, particularly in the context of transcriptome and chromatin accessibility. Here, scRNA-seq and scATAC-seq methods were employed to investigate the role of ECs involved in SSC-ILD in order to better understand cellular heterogeneity at the single-cell multiomics level. Furthermore, we used the scATAC-seq with SSC genome-wide association studies (GWAS) loci to elucidate mechanisms for functional GWAS loci.

## Methods

### Data download

scRNA-seq for 11 individuals and matched scATAC-seq for 5 individuals were conducted in the recent study [4]. Therefore, all these raw fastq files were downloaded from the Gene Expression Omnibus (accession number: GSE212109)

### scATAC-seq processing

scATAC-seq data from the scATAC-seq experiment were aligned to the GRCh19 (hg19) reference genome and quantified using the Cellranger-atac count tool (10× Genomics, v.2.0). ArchR v1.0.2 [10] was used to construct arrow files by reading in the available read fragments for each sample and applying the default augments. We selected cells with less than 1,000 unique fragments and enrichment at TSSs below 4 to ensure that each cell had a strong signal and was well-sequenced. Meanwhile, doublets were inferred and filtered by ArchR. The method scOpen (v.1.0.0) [11] was used to construct a low-dimensional matrix of the cells for dimensionality reduction. To correct the batch effects (sample and chemistry) and integrate the data, the Harmony (v.0.1.1) algorithm [12] was used, and cells were clustered using the Leiden algorithm with a resolution of 1, respectively. A gene activity score matrix was built using the method “addGeneScoreMatrix” to annotate the clusters and marker genes were discovered for each cluster using the function “getMarkerFeatures”. We also utilized the MAGIC [13] imputed weight approach on the obtained gene activity scores to reduce noise in the data scarcity in scATAC-seq. The clusters were annotated using the same markers from the scRNA-seq data. Clustering with “FindClusters” with a default resolution of 0.5 during the reclustering process to better find small clusters. In two-dimensional space, all data were represented using uniform manifold approximation and projection (UMAP). MACS2 was utilized to accomplish peak calling [14], and the peak matrix was then generated using “addReproduciblePeakSet” and these pseudobulk duplicates. Non-overlapping pseudobulk replicates were constructed from groups of cells using the “addGroupCoverages” function with distinct inputs for differential comparisons of cell types and clinical stages. The pseudobulk peak set was subjected to motif enrichment and motif deviation studies. The chromVAR [15] deviation scores for these motifs were also estimated using the ArchR implementation. Using the “getMarkerFeatures” function, the pseudobulked peak set was employed for differential analysis between different cell types and disease states. To calculate the *p*-value and false discovery rate (FDR) between any pair of samples, the Wilcoxon test and the Benjamini-Hochberg (BH) multiple tests were utilized. FDR<=0.01 and log2-fold change (FC)>=1 were used to determine differentially accessible distal peaks. Meanwhile, we used the ChIPseeker package’s “annotatePeak” function [16] to annotate the closest genes in the peak region using default inputs. Tn5 insertions in genome-wide motifs were measured and normalized by eliminating the Tn5 bias from the footprinting signal. We normalized these footprints using mean values 200-250 from the motif center. To identify peak-to-gene links, we used the ArchR “addPeak2GeneLinks” function and set the parameters to “corCutOff=0.45” and “resolution=10,000”, with “reducedDims” as the dimensionality reduction results after batch correction.

### Overlap of SSC GWAS SNPs with cell-type peaks

We downloaded summary statistics from López-Isac E *et al* [17] who performed GWAS meta-analysis in 26,679 European populations involving 9,095 cases and 17,584 controls for comparison with GWASs of SSC via GWAS catalog (https://www.ebi.ac.uk/gwas) (accession codes GCST009131). The same strategy as described by Turner *et al* was used to detect overlap between disease-related SNPs and cell-type peaks [18]. In brief, the lead variants were utilized as input to PLINK (version 1.9) [19] first, and then kept variants with r^2^>0.8 utilizing the European population. To accommodate for SSC variants located close to a peak, a 100-bp window with the SNP in the center was evaluated by extending the SNP location 50 bp in each direction using BEDOPS (v.2.4.37) [20]. Finally, bedtools (v.2.26.0) [21] was used to intersect these SNP with each cell type peak.

### scRNA-seq processing

For each sample, the raw fastq files were processed and aligned to the GRCh38 reference using CellRanger software with the default parameters (https://support.10xgenomics.com, version 7.1.0). Following that, the Seurat R package v4.2.0 [22] was used for data quality control, preprocessing, and dimensional reduction analysis, as previously described [23]. After generating gene-cell data matrices from combined datasets, matrices were merged, and poor-quality cells with 200 or more expressed genes and mitochondrial gene percentages greater than 50 were excluded, leaving 48,622 cells for further analysis. Doublets were detected using the Scrublet package (version 2.0.2) using the “scrub.scrub_doublets()” function [24] and removed by Seurat. After normalizing the data using log transformation and scaling, the Harmony R package v1.0 was used to correct batch effects (sample and chemistry). For downstream UMAP visualization and clustering, the top 30 dimensions of Harmony embeddings were used. Cells were clustered using the Louvain algorithm with a resolution of 0.2, yielding 16 unique cell clusters. We utilized the Seurat package’s “FindAllMarkers” function with the default parameters for scRNA-seq differential expression analysis. A BH-adjusted *p*-value of less than 0.05 was regarded as statistically significant. CellTypist [25] was used to determine the cell type annotations for each cluster based on the expression patterns of known marker genes.

### Multiomics data processing

An anchoring strategy as implemented in the Archr packages was employed to carry out an integrated analysis of scATAC-seq and scRNA-seq data. Briefly, a collection of “anchors” between datasets presumed to be in a comparable biological state was first found (the “FindTransferAnchors” function) using canonical-correlation analysis (CCA). These “anchors” are then utilized as a reference to integrate the datasets, and the cluster labels from the scRNA-seq data can be projected onto the scATAC-seq data using the “TransferData” function. To assess the agreement between annotated and predicted cell labels in scATAC-seq data, an adjusted rand index was computed.

### Pseudotime and RNA velocity analysis

In the scATAC-seq and integrated datasets, we created a differentiation trajectory and aligned single cells along it. Using batch-corrected Harmony embeddings, a cellular trajectory was built in a low-dimension space based on the trajectory backbone. The cells were assigned based on pseudotime value estimates, and differential feature z-scores were used to generate a heatmap. For pseudotime trajectory analysis in scRNA-seq, R packages Monocle3 (version 1.3.1) [26] were utilized with their default parameters. The root point was determined as “EC venous systemic”. For RNA velocity analysis, CellRanger produced BAM files in conjunction with velocyto were used to generate a loom file containing the quantification of spliced and unspliced RNA [27]. Following that, we concatenated samples and computed RNA velocity using the packages scvelo with the default settings, as previously described [23].

### Receptor-ligand pair analysis

The Python software CellPhoneDB [28] with database version 4 was used to infer cell-cell communication networks from single-cell transcriptome data to measure cellular crosstalk across various cell types. Only receptors and ligands expressed in more than 5% of the cells in a given cell type were taken into account. We retained only statistically significant averages (*p*<0.05) of interaction partners from the CellPhoneDB output, which served as a proxy for mean expression of both the ligand and the receptor of a given anticipated relationship.

### Identifying TF target genes

To identify significantly shared TFs and their directly regulated target genes in SSC-ILD disease, we used the method described by Kuppe *et al* [29], with minor modifications. In brief, following pseudotime analysis, the transcription factors and RNA expression along the trajectory were sorted and a pseudotime label for each transcription factor/gene was assigned. To associate the selected TF with targets, we explored the correlation of peak accessibility and gene expression to identify peak-to-gene links and further subset these links by those containing the TF motif. Significantly correlated links (FDR<0.0001) with a positive correlation were selected. To build a quantitative transcription factor-gene-regulatory network, we estimated the correlation of the transcription factor binding activity between scATAC-seq and target gene expression from scRNA-seq data, and only those interactions with a Pearson correlation >0.8 were considered.

## Results and Discussion

### Single-cell landscape in systemic sclerosis-associated interstitial lung disease

A total of 48,622 cells were investigated using scRNA-seq data (**Supplemental Figure 1A**), including 18,591 cells from 6 healthy individuals and 30,031 cells from 5 SSC-ILD patients, while 2 samples from SSC-ILD and the remaining 3 samples were included in scATAC-seq (**Supplemental Figure 1B**). Following that, batch adjustment for samples and chemistry was conducted using the R package “Harmony” on both the scATAC-seq and scRNA-seq datasets. Based on the expression of lineage-specific markers, 15 main cell types in the lung were identified in scRNA-seq (**Figure 1A**-**1C**). In detail, macrophages, monocytes, T cells, monocyte-derived Mph, multiciliated, fibroblasts, ECs, basal resting, NK cells, alveolar type 2 epithelial cells (AT2), mast cells, smooth muscle, AT1, B cells, lymphatic EC mature, and an unidentified cell population were reported (**Figure 1C**). We obtained a total of 14,717 high-quality cells after filtering for scATAC-seq and identified 13 clusters in ArchR (**Figure 1D**-**1E**). 12 cell types were annotated based on gene activity scores provided to each cluster (**Supplemental Figure 1C**). Label transfer was accomplished by generating a gene-activity matrix from scATAC-seq data, which is a measurement of chromatin accessibility inside the gene body and promoter of protein-coding genes. Transfer anchors have been identified between the “reference” scRNA-seq datasets and the “query” gene activity matrix, and anticipated cell types were assigned. When comparing the annotated label using the scRNA-seq datasets, the 12 cell types identified for scATAC-seq showed high consistency, demonstrating that scATAC-seq is equivalent to scRNA-seq in the detection and assignment of cell identities (**Figure 1F**). Following that, the MACS2 was used to evaluate the differences in chromatin accessibility (DAR) between cell types. A master set of 50,229 peaks was generated by using a log2-fold-change threshold of 1 and adjusted *p*-values to measure DAR significance (FDR<=0.01). Furthermore, 26% of DAR were found at a promoter region less than 3 kb from the nearest transcriptional start point (**Figure 1G**). Intronic was the second most common site, and DAR distribution was relatively consistent across cell types. Meanwhile, motif enrichment analysis for these marker peaks was undertaken to identify the TFs likely driving the regulatory profiles in each cell type (**Figure 1H**). Top enriched motifs in T cells (ERG, ELK4, and ERF) were discovered. Furthermore, SOX13 motifs were shown to be enriched in ECs, as well as ZNF238 and ZBTB42 motifs in fibroblasts. Overall, the integrated datasets allows us to detect cell type heterogeneity at the transcriptional and chromatin accessibility levels (**Figure 1I**).

**Figure 1.**
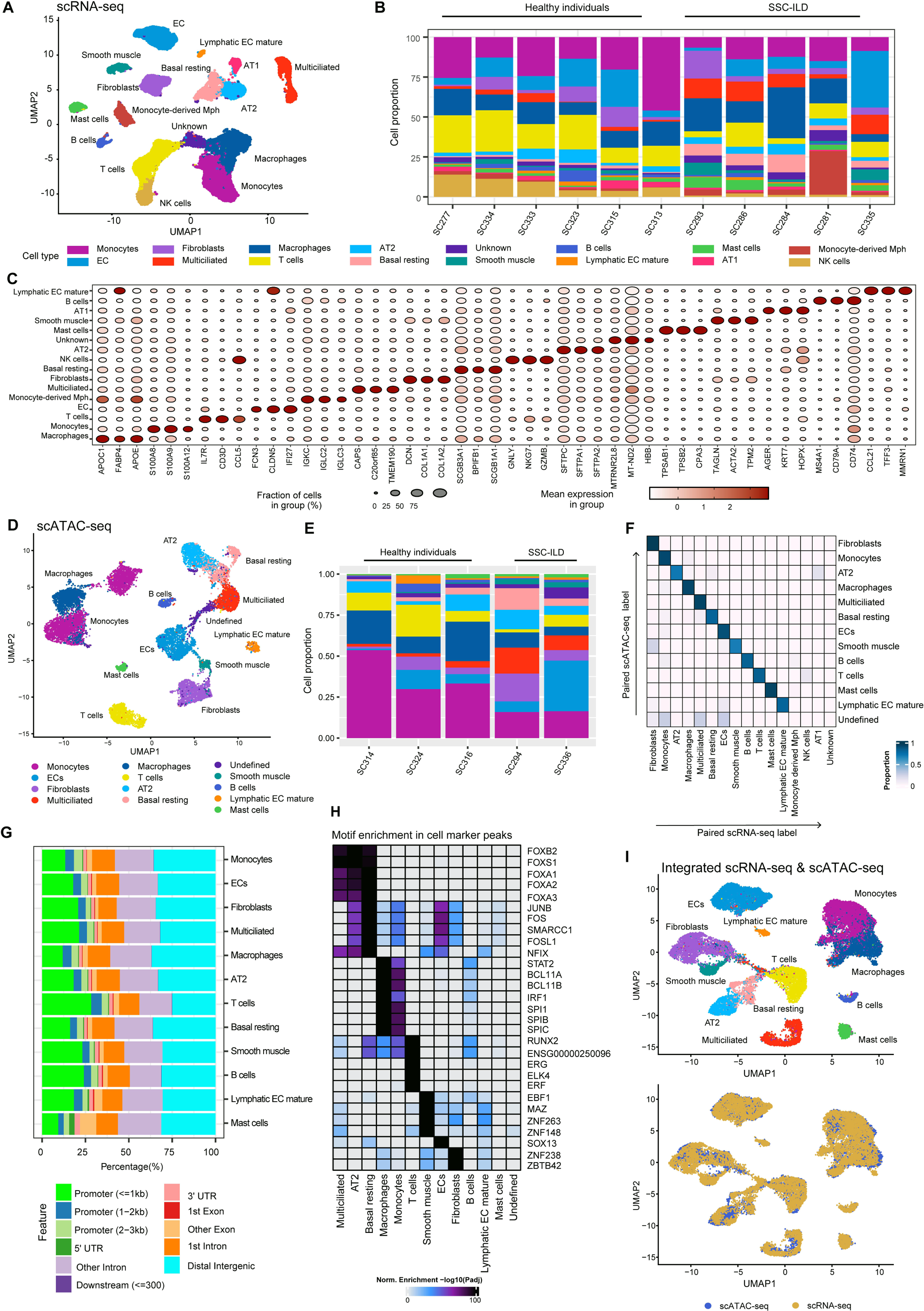
An overview of the epigenetic and transcriptional landscape of cell types in the lungs of SSC-ILD patients and healthy individuals. **(A)** UMAP projection of 48,622 scRNA-seq profiles of cell types in the lung from 11 individuals. Individual cells are represented by dots, and different cell types are indicated by different colors. **(B)** Dot plots of gene expression of the marker genes in the scRNA-seq datasets. The size of the dots represents the percentage of cells in each cluster that have the gene of interest. Color intensity denoted the standardized gene expression level. **(C)** Barplots depicting the cell-type proportions of scRNA-seq data across all samples. The various colors represent the major cell types. **(D)** UMAP projection of 14,717 scATAC-seq profiles of lung cell types from 5 individuals. Individual cells are represented by dots, while different cell types are represented by different colors. **(E)** Barplots of scATAC-seq cell-type proportions across all samples. The colors represent the several major cell types. **(F)** Validation of the scATAC-seq cell type annotation using paired 5 scRNA-seq data. Heatmap displaying evaluation findings based on the adjusted rand index (ARI). **(G)** Annotated DAR location bar plot for each cell type. **(H)** Heatmap of TF motifs found in peak sequences of cell-type markers. The color indicates the normalized motif enrichment score in ArchR obtained with HOMER and the hypergeometric test. **(I)** Multi-omics integration for scATAC-seq and scRNA-seq data.

### Heterogeneity of endothelial cell subsets

Several investigations have demonstrated that ECs have a role in the pathophysiology of SSC [30–31]. The chromatin accessibility of ECs from control and SSC-ILD lungs, however, remains unclear. To investigate the heterogeneity of ECs subsets in SSC-ILD, we first examined the differences in ECs subsets between HC and SSC-ILD patients in scRNA-seq including 11 individuals due to the higher statistical power. Following the re-clustered ECs (4,492 cells), 5 cell subpopulations were identified based on particular gene expression (**Figure 2A** and **Supplemental Figure 2A-2C**). Interestingly, our study identified two cell subpopulations of ECs with unequal frequencies in SSC-ILD patients and healthy individuals (*p*<0.01) (**Figure 2B** and **Supplemental Figure 2D**). The frequency of SSC-ILD was found to be higher in EC venous systemic, which substantially expressed RBP7; however, the frequency of SSC-ILD was significantly lower in EC aerocyte capillary, which highly expressed TBX2. Our findings from RNA velocity and pseudotime analysis utilizing scRNA-seq revealed the differentiation of EC venous systemic to EC aerocyte capillary, indicating that EC venous systemic could serve as a precursor stages for EC aerocyte capillary (**Supplemental Figure 2E-2F**). Moreover, the gene ontology analysis of EC venous systemic derived from patients with SSC-ILD were mainly enriched in the response to unfolded protein, regulation of Wnt signaling pathway, and positive regulation of inflammatory response (**Supplemental Figure 2G-2H**). These results suggested that EC venous systemic in SSC-ILD patients could be generally activated and proinflammatory.

**Figure 2.**
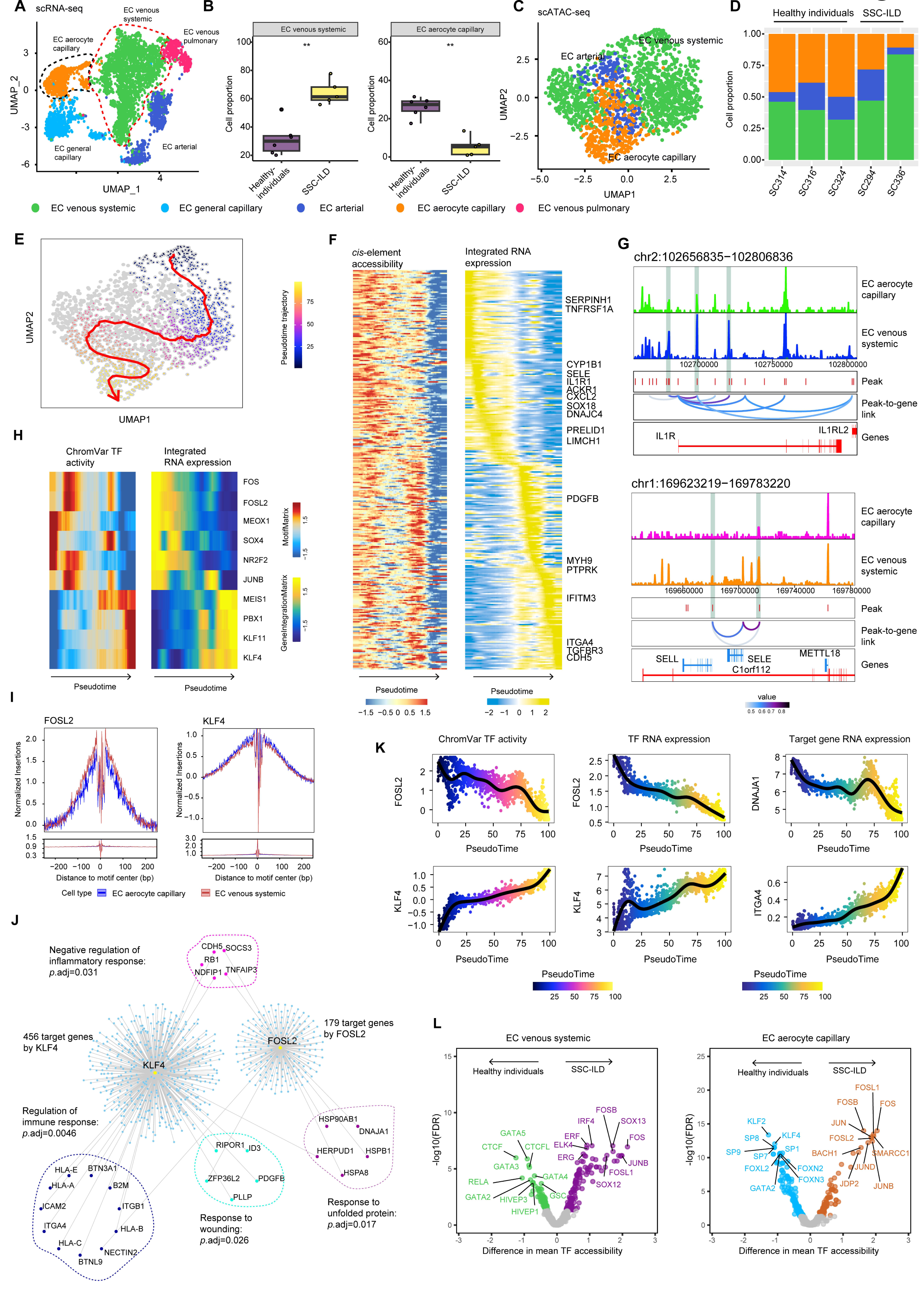
Epigenomic and transcriptional signatures of EC subsets. (**A**) UMAP projection of 4,492 EC subpopulation scRNA-seq profiles. Individual cells are represented by dots, while different cell types are represented by different colors. (**B**) Differences in chosen population proportions across healthy individuals (n=6) and SSC-ILD groups (n=5). The Wilcoxon test was used to get the *p*-values. (**C**) UMAP projection of 1,760 scATAC-seq profiles of lung cell types from 5 individuals. Individual cells are represented by dots, while different cell types are represented by different colors. (**D**) Barplots of scATAC-seq data cell-type proportions across all samples. The colors represent the indicated cell types. (**E**) UMAP depicting the ECs lineage trajectory. Pseudotime values were superimposed on the UMAP embedding; the smoothed line and arrow indicate the representation of the spline fit trajectory route. (**F**) Heatmaps depicting the 323 ordered gene-score and matched RNA expression trajectory from EC venous systemic to EC aerocyte capillary across pseudotime. (**G**) Accessibility tracks surrounding the IL1R (up) and SELE (down) loci. (**H**) Heatmaps of 10 positive TF regulators whose TF motif activity (left) and corresponding gene expression (right) are positively associated across the ECs differentiation pseudotime trajectory in (E). (**I**) Comparison of aggregate TF footprints for FOSL2 and KLF4 in EC venous systemic and EC aerocyte capillary, respectively. (**J**) A TF regulatory network depicting the TFs (FOSL2 and KLF4) and probable target genes. (**K**) chromVAR bias-corrected deviation scores, RNA expression, and selected target gene expression across ECs pseudotime for the specified TFs. In an individual pseudotime-ordered scATAC-seq profile, each dot indicates the deviation score or RNA expression. The smoothed fit spanning pseudotime and chromVAR deviation scores or RNA expression is represented by the line. (**L**) Volcano plots exhibiting the differential TF motif accessibility in the chromVAR TF bias-corrected deviation between health and SSC-ILD individuals in the identified ECs subpopulations using the mean TF motif accessibility. The *p*-values were obtained using a Wilcoxon test, and the FDR was performed using the Benjamini-Hochberg method.

The integrated analysis of scRNA-seq and scATAC-seq was then performed to better understand the chromatin accessibility of ECs subpopulations (**Figure 2C**-**2D**). Label transfer from scRNA-seq to scATAC-seq revealed the three cell types, with a consistent lower frequency for EC venous systemic and a greater frequency for EC aerocyte capillary, as expected (**Figure 2C**-**2D** and **Supplemental Figure 2I-2J**). A pseudotime analysis was undertaken to uncover the dynamics and modulation of cell fate decisions in EC subpopulations (**Figure 2E**). A lineage route from EC venous systemic to EC aerocyte capillary was discovered by pseudotime analysis utilizing scATAC-seq. The examination of *cis*-elements near certain genes and the accessibility of TF motifs revealed distinct regulatory patterns of accessibility over the pseudotime (**Figure 2F**-**2H**). Interestingly, *cis*-element accessibility in some loci involved in inflammatory response, such as IL1R1 and SELE, as well as response to unfolded protein, such as SERPINH1 and DNAJB1, gradually decreased from EC venous systemic to EC aerocyte capillary, demonstrating that the expression of inflammatory response-related genes was accurately regulated by a distinct *cis*-element network in the different states (**Figure 2F**-**2G**). The distinct TF activities across the pseudotime of EC venous systemic to EC aerocyte capillary paths were revealed by integrating chromVAR TF-activity and TF RNA expression studies (**Figure 2H**). For example, by assigning a path of accessibility from differentiated EC venous systemic to EC aerocyte capillary, we identified enriched MEOX1 motifs at the start of the trajectory, followed by enrichment of FOS, FOSL2, and JUNB motifs in EC aerocyte capillary, and finally PBX1, KLF11, and KLF4 motifs. According to the trajectory analysis, the EC venous systemic showed significantly higher occupancy in FOSL2 in the footprint analysis than the EC aerocyte capillary, while the EC aerocyte capillary showed a more pronounced DNA occupancy of KLF4, implying a role for FOSL2 and KLF4 as distinct transcription factors in shaping EC maturation (**Figure 2I**). We used a recently described approach to identify probable TF target genes based on the merged scATAC-seq and scRNA-seq datasets to demonstrate the TFs regulatory program network in the ECs [28]. Only Pearson correlations greater than 0.8 between transcription factor binding activity from scATAC-seq and target gene expression from scRNA-seq data were evaluated. Using this method, we discovered 456 genes controlled by KLF4 and 179 genes regulated by FOSL2 (**Figure 2J**). For example, four genes implicated in the response to unfolded protein, namely HSPA8, HSP90AB1, DNAJA1, and HSPB1, were shown to be regulated by FOSL2, which likewise showed decreased activity along the pseudotime axis (**Figure 2K**). Furthermore, our analysis found that KLF4 regulates the expression of genes involved in immune response regulation, such as BTNL9, ITGA4, and ICAM2, in EC aerocyte capillaries. In the pseudotime analysis, we found increased KLF4 binding activity and SITGA4 expression, supporting the significance of KLF4 as a possible activator of immune response activation (**Figure 2K**). The chromVAR motif variations in the EC venous systemic and EC aerocyte capillary between healthy individuals and SSC-ILD patients were also compared (**Figure 2L**). AP-1 factors, including FOS and JUNB, were shown to be abundant in both EC venous systemic and EC aerocyte capillary of SSC-ILD patients. Furthermore, transcription factors including as GATA2 and GATA3 that are involved in the negative regulation of ECs apoptosis were shown to be abundant in the EC venous systemic of healthy individuals. Overall, our analysis indicated epigenetic reprogramming in the formation of ECs, with activation characteristics and compositional and epigenomic abnormalities of EC venous systemic in SSC-ILD patients.

### Annotation of target cell types at SSC GWAS loci

Indeed, the paired scATAC-seq/scRNA-seq datasets enabled us to deconstruct the mechanisms underlying previously reported causative risk variations from GWASs and to uncover disease-relevant cell types associated with these loci. Several studies have found that noncoding GWAS variants are concentrated in CREs and frequently function in cell-type-specific ways [32–34]. In this study, a multi-tiered strategy was employed to prioritize putative functional SSC GWAS variants, as described by Turner *et al* [18]. In brief, after identifying CRE variants, we compared SSC lead variants (and variants in high LD (r2>0.8; EUR)) from the most recent SSC GWAS meta-analyses in a European cohort of 26,679 individuals with scATAC peaks (**Figure 3A**). This identified a subset of variants (50 bp) that overlapped GWASs loci and marker peaks, with the majority of SSC SNPs located within macrophage peaks, followed by monocytes, T cells, AT2/basal resting cells, and B cells peaks (**Figure 3B**). We next identified target cell types for SSC regulatory variants based on these overlaps. Several top potential SSC variants, for example, map to cell-type-specific peaks, such as rs3792783 at the TNIP1 locus within an AT2 and basal resting cells peak and rs1005714 at the NUP85 locus within macrophages (**Figure 3C**). Other SSC variants, such as rs2305743 at IL12RB1 and rs1378942 at MIR4513, map to peaks shared by T cells, monocytes, and macrophages. We reclustered AT2 using a scRNA-seq datasets of 11 individuals to characterize its role in SSC-ILD, and four clusters were identified (**Supplemental Figure 3A-3C**). Notably, the frequencies of cluster 3, which highly expressed SERPINF1, were considerably greater in patients with SSC-ILD (**Supplemental Figure 3D**). RNA velocity analysis suggests a C1 to C3 lineage trajectory (**Supplemental Figure 3E**). Due to the small number of cells observed in scRNA-seq for AT2 cluster 3, which may represent a rare cell population, scATAC-seq did not disclose this subpopulation (data not shown). Furthermore, because of the small number of samples included in the study, no substantial overlap with GWAS loci was discovered for the cell-type peaks of fibroblasts, one of the main effector cells in the extracellular matrix remodelling of the disease [3]. A larger datasets should be used in the future to explore these subpopulations in depth.

**Figure 3.**
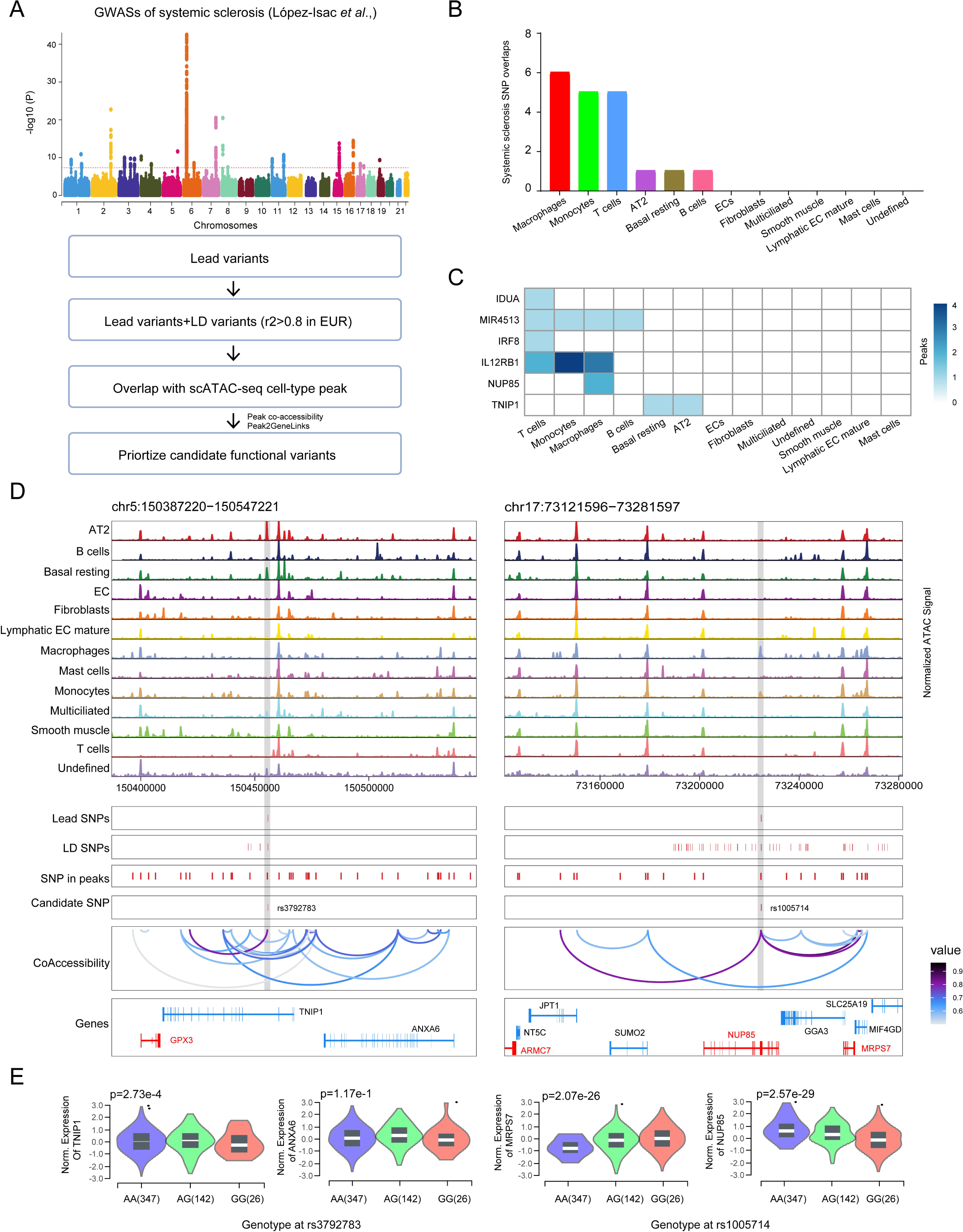
scATAC-seq identifies mechanisms for functional SSC GWAS variants. (**A**) Prioritizing potential SSC-associated GWAS SNPs using a multi-tiered method, as described by Turner *et al*. (**B**) LD-expanded (r^2^>0.8; EUR) SSC GWAS variants (50 bp) overlap with cell-type peaks. (**C**) Heatmap displaying the number of peaks per cell type that intersect SSC GWAS loci. (**D**) Normalized chromatin accessibility landscape for cell-type-specific pseudobulk tracks in the vicinity of the TNIP1 (left) and NUP85 (right) loci. Below the scATAC-seq tracks are the positions of the scATAC-seq peaks, the GWAS lead SNP, the SNP candidates in LD with the lead SNP, and the candidate functional SNP. (**E**) Lung eQTL in GTEx for rs3792783 and rs1005714. The G allele for rs3792783 is strongly related to lower TNIP1 mRNA levels in GTEx lung (left two panels). The G allele for rs1005714 is strongly related to elevated MRPS7 and NUP85 mRNA levels (right two panels). The eQTL *p*-value is displayed as a result of the GTEx pipeline’s linear regression between genotype and normalized gene expression levels. The boxplot within the violin plot includes the median and interquartile range (IQR) from 25% to 75%.

Because noncoding variants cannot always regulate the adjacent gene, we used co-accessibility and scRNA-seq integration to link candidate variants to target gene expression (**Supplemental Figure 4A**). When all cell types were aggregated, this orthogonal method yielded a total of 31,432 Peak2Gene linkages (**Supplemental Figure 4A**). It is worth noting that the macrophages peak-containing variant, rs1005714, has a co-accessibility and gene expression relationship with MRPS7 (**Supplemental Figure 4B**), and is also an MRPS7 lung-specific expression quantitative trait locus (eQTL) in the Genotype-Tissue Expression project (GTEx) (**Figure 3E**). MRSPS7, but not NUP85, is differently expressed between healthy individuals and SSC-ILD patients (uncorrected *p*=2.9e-05) (**Supplemental Figure 4C**). These results suggested that MRSPS7 could be a candidate gene associated with SSC, a larger datasets as well as experiments should be performed to validate these results.

### Cell-cell communication network

To identify the reciprocal communication between different cell types in the lung, we investigated the accumulated ligand/receptor interaction database CellPhoneDB to detect reciprocal communication. The number of cell-cell interactions was increased in SSC-ILD patients than in healthy individuals, as expected (**Figure 4A**). Fibroblasts, in particular, demonstrated the greatest capacity for cell-cell interactions and shared an increased number of projected connections with lymphatic EC mature, ECs, and fibroblasts in patients with SSC-ILD. Furthermore, with the exception of monocyte-derived Mph, ECs increased cell-cell interactions with practically all other cell types in SSC-ILD patients. These findings further suggest that ECs may play an essential role in disease progression.

**Figure 4.**
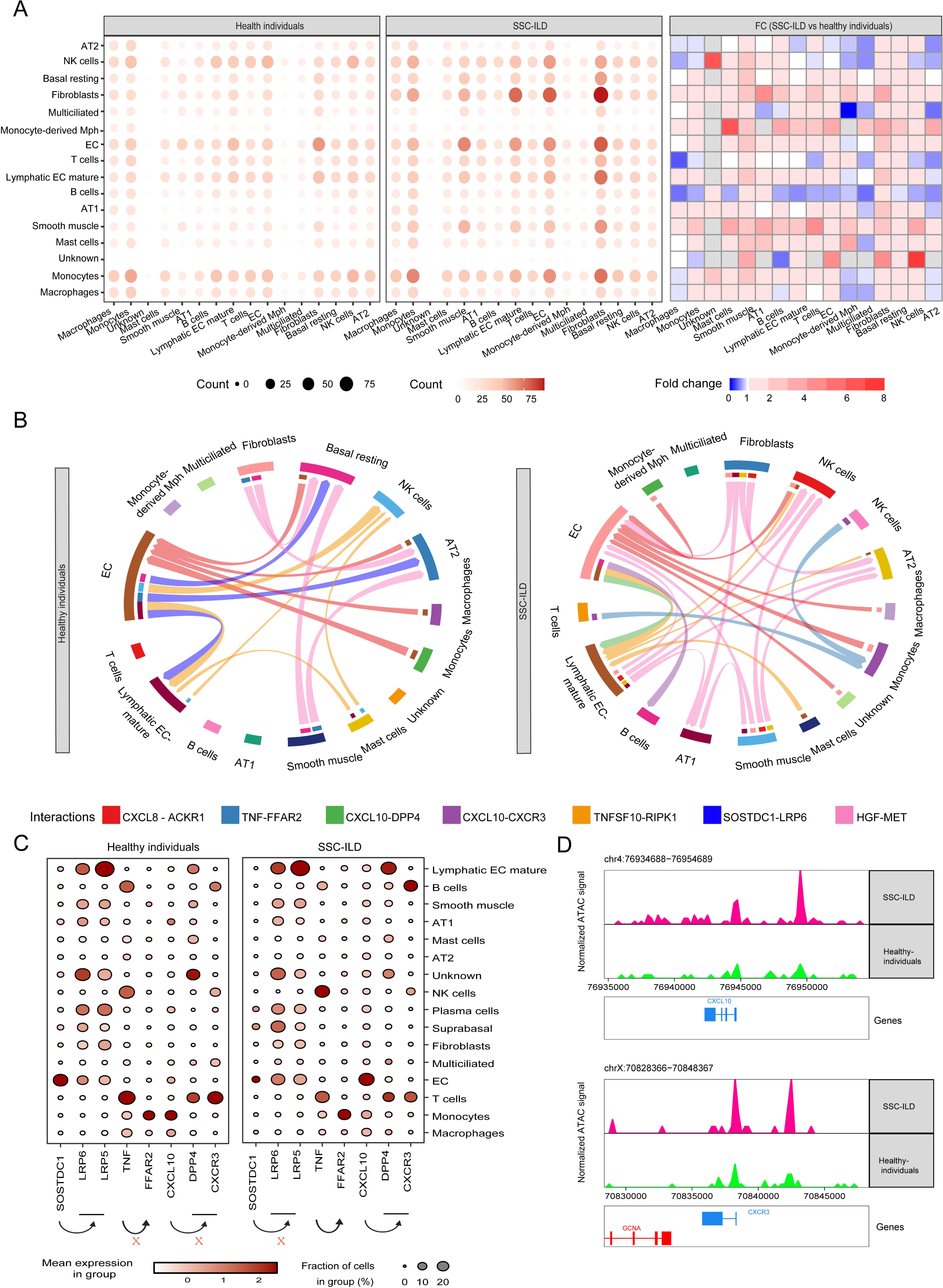
The cellular-cellular communications of cell types in lung. (**A**) A dotplot depicting the differences in ligand-receptor interaction events between cell types in the healthy and SSC-ILD groups. The dotplot (left) and heatmap (right) show the number of interactions and fold changes (SSC-ILD vs healthy individuals). (**B**) Circos plots depicting genes implicated in ligand-receptor interactions between cell types. Color scale is proportional to interaction strength as assessed by CellPhoneDB interaction means of statistically significant interactions. (**C**) Dot plots demonstrating an increase in gene expression for the specified genes. The size of the dots represents the percentage of cells that express the marker. The color scale shows the average gene expression levels. The arrows represent ligand-receptor pairings generated from CellPhoneDB. The X represents signaling that has been disabled. (**D**) Single-cell chromatin accessibility in the CXCL10 and CXCR3 loci as seen by genome browser tracks.

In SSC-ILD patients, ECs had enhanced CXCL10-DPP4 and CXCL10-CXCR3 interactions with lymphatic EC mature and B cells (**Figure 4B**). According to gene ontology enrichment analysis, CXCL10 was substantially enriched in the RIG-I-like receptor signaling pathway and ECs activation (*p.*adj<0.01), whereas CXCL10 and CXCR3 were significantly involved in the inflammatory response (*p*.adj=0.00081). CXCL10 and its receptors CXCR3 were shown to be more accessible in SSC-ILD patients, with increased RNA expression of CXCL10 and its receptor CXCR3 (**Figure 4C**-**4D**). Interestingly, TNFSF10-RIPK1 and TNF-FFAR2 signaling was anticipated to be active during AT2 and monocyte interactions with other cell types in SSC-ILD patients. Furthermore, SOSTDC1-LRP6 interactions were expected to be reduced in the communication between EC cells with basal resting cells and AT2 in SSC-ILD patients (**Figure 4B**). In fact, this has been identified as a critical regulatory interaction in the negative control of the canonical Wnt signaling pathway [35]. Overall, our findings point to a complicated interaction network between healthy individuals and patients. More research is needed to determine whether ECs are involved in the regulation that leads to the onset of the inflammatory response in B cells.

In conclusion, by integrating single-cell multiomics data, our research demonstrated that systemic-venous ECs may play a significant pro-inflammatory role in SSC patients, providing insights into the pathophysiology of SSC and therapeutic alternatives.

## Supporting information

Supplementary Table 1. The statistics generated in the study.

## Data Availability

All data produced in the present work are contained in the manuscript.

## Acknowledgments

The authors would like to thank Anna and her colleagues for their nice work and the generation of the single-cell multiomics datasets.

## Figure Legend

**Figure S1.**
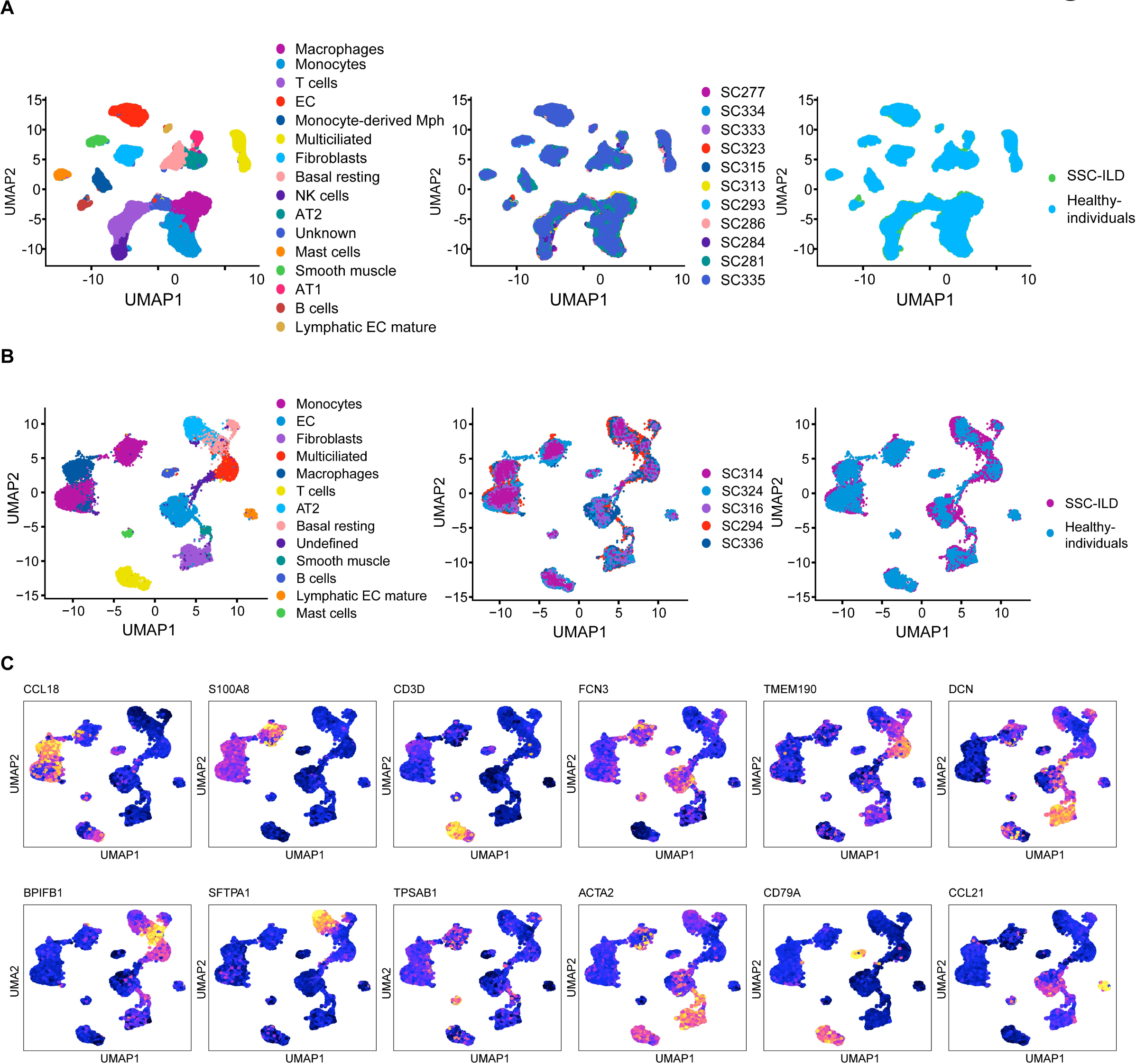
shows an overview of the single-cell RNA and ATAC datasets. (**A**) UMAP embedding of scRNA-seq data from all samples for cell types, samples, and disease states. (**B**) UMAP embedding of scATAC-seq data from all samples for cell types, samples, and disease states. CCL18 (macrophages), S100A8 (monocytes), CD3D (T cells), FCN3 (endothelial cells), TMEM190 (multiciliated), DCN (fibroblasts), BPIFB1 (basal resting), SFTPA1 (AT2), TPSAB1 (mast cells), ACTA2 (smooth muscle), CD79A (B cells), and CCL21 (lymphatic EC mature).

**Figure S2.**
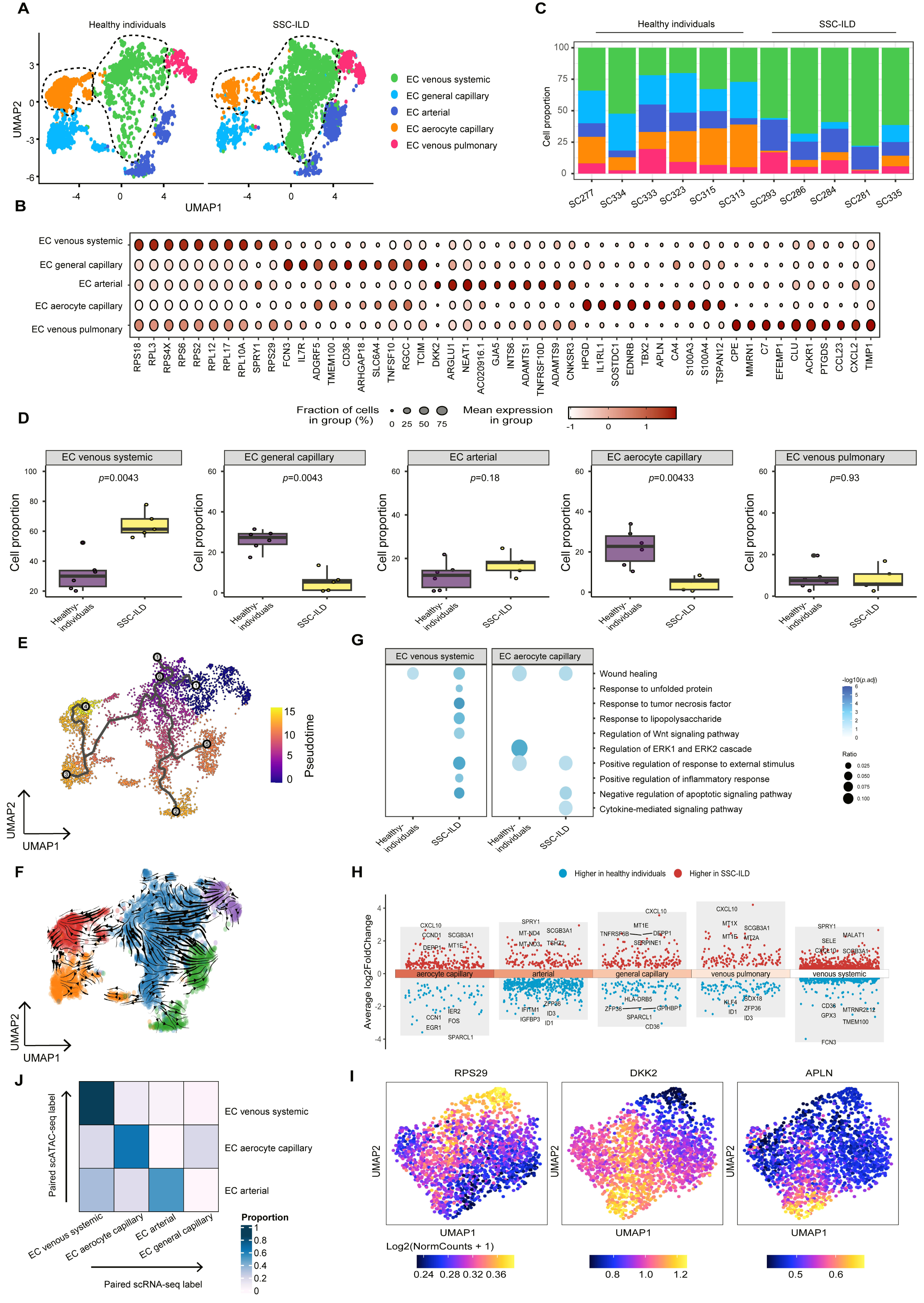
Sub-clustering of ECs and integration of scRNA-seq and scATAC-seq datasets. (**A**) UMAP embedding of scRNA-seq data from 11 samples in the SSC group and health individuals. (**B**) Dot plots showing marker gene expression in the scRNA-seq datasets. The dot size represents the percentage of cells in each cluster that have the gene of interest. The color intensity reflected the standardized gene expression level. (**C**) Barplots of scRNA-seq data cell-type proportions across all samples. The colors represent the several primary cell kinds. (**D**) Proportions of EC subpopulations differ between healthy individuals (n=6) and SSC-ILD groups (n=5). The Wilcoxon test was used to compute the *p*-values. (**E**) Monocle 3 inferred scRNA-seq EC venous systemic differentiation route towards EC aerocyte capillary. Pseudotime colors the cells. (**F**) RNA velocity cell trajectory analysis of ECs using scRNA-seq. (**G**) GO enrichment of distinctly expressed genes in EC venous systemic and EC aerocyte capillary between healthy individuals and SSC-ILD patients. (**H**) Differential gene analysis of significantly upregulated and downregulated genes between healthy individuals and SSC-ILD patients across cell types. The Y-axis denotes the average log2FoldChange. red: upregulated in SSC-ILD; blue: downregulated genes in SSC-ILD. (**I**) UMAP plot of scATAC-seq datasets by gene score for cell-type marker genes, including RPS29 (EC venous systemic), DKK2 (EC arterial), and APLN (EC aerocyte capillaries). (**J**) Validation of scATAC-seq cell type annotation using scRNA-seq data. Heatmap showing evaluation results by adjusted rand index (ARI).

**Figure S3.**
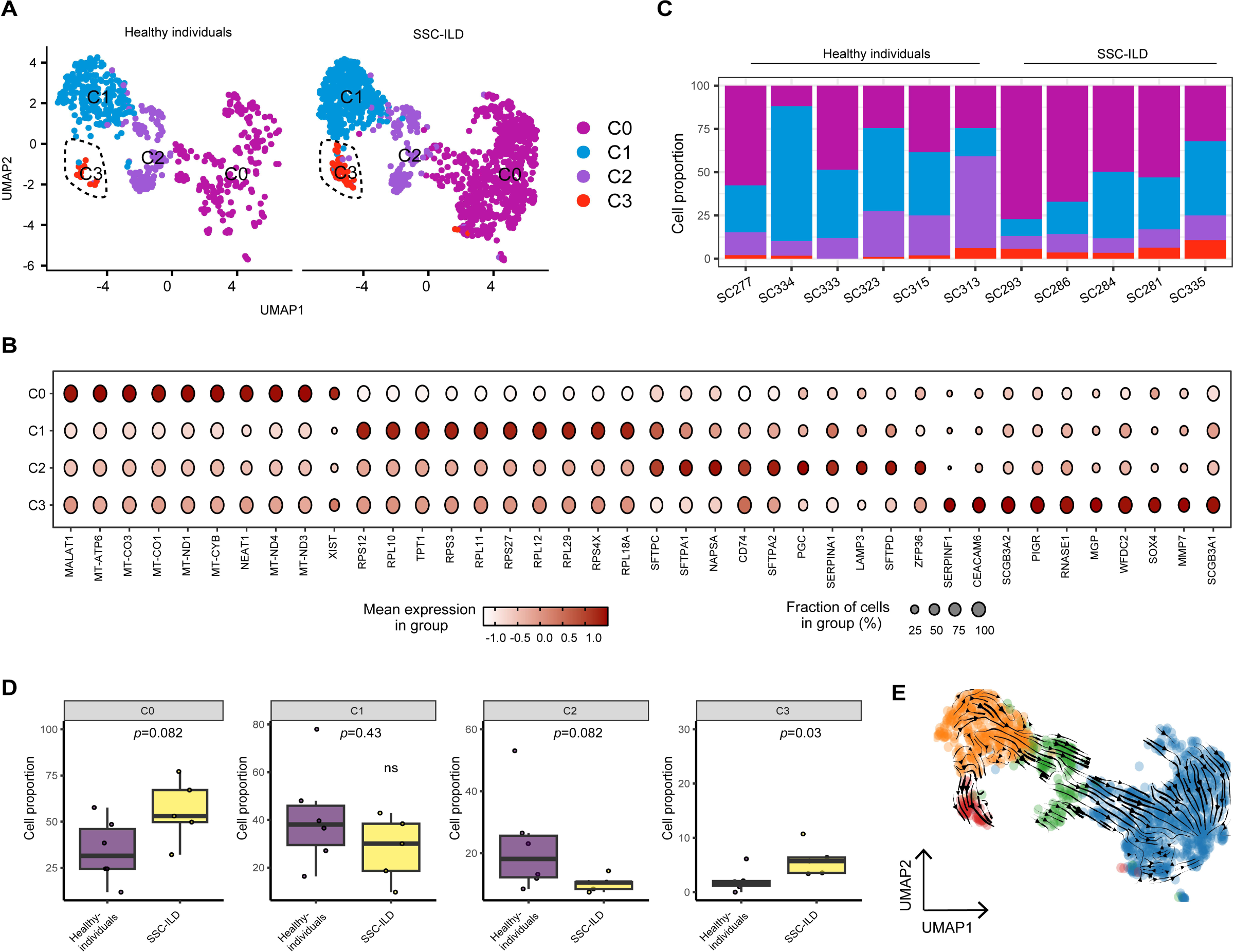
Sub-clustering of AT2 using scRNA datasets. (**A**) UMAP embedding of scRNA-seq data from 11 SSC and healthy individuals. (**B**) Dot plots showing marker gene expression in the scRNA-seq datasets. The dot size represents the percentage of cells in each cluster that have the gene of interest. The color intensity reflected the standardized gene expression level. (**C**) Barplots of scRNA-seq data cell-type proportions across all samples. The colors represent the several primary cell kinds. (**D**) Differences in the proportions of four AT2 clusters among healthy individuals (n=6) and SSC-ILD groups (n=5). The Wilcoxon test was used to compute the *p*-values. (**E**) AT2 RNA velocity cell trajectory analysis. AT2, alveolar type 2 epithelial cell.

**Figure S4.**
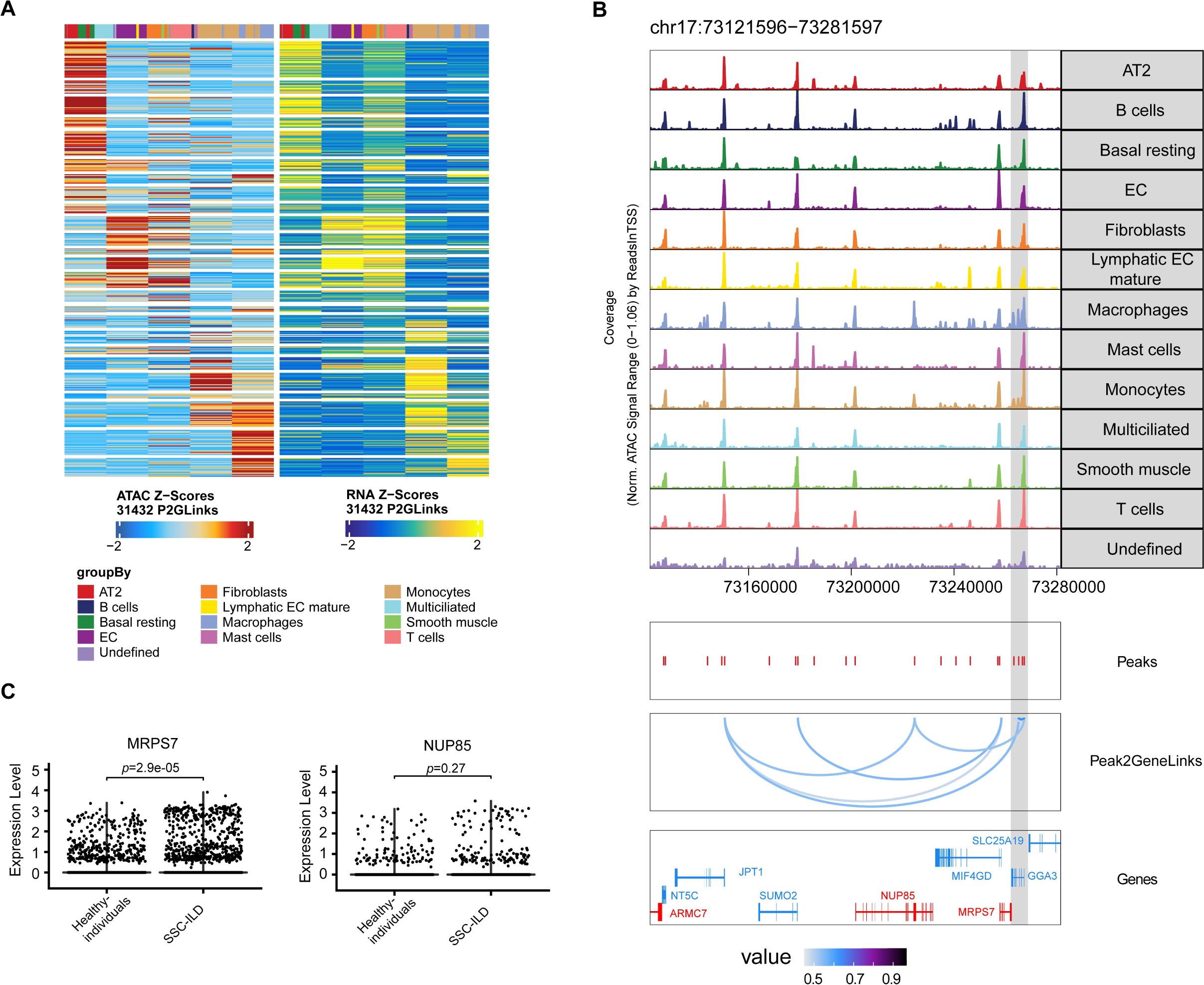
Co-accessibility of scATAC-seq and integration with scRNA-seq link putative regulatory elements to target gene. (**A**) Heatmap summary of ArchR Peak2Gene connections (n=31,432) at a resolution of 10 kb, where chromatin accessibility is substantially linked with target gene expression. The Z-scores for scATAC-seq peak accessibility are shown on the left, while the Z-scores for RNA expression are shown on the right. (**B**) *cis*-regulatory architecture and lung cell types at the MRPS7 GWAS loci. (**C**) Violin plot of gene expression in the indicated illness stages. Each dot represents the expression of a single gene in a single cell.

**Supplementary Table 1. The statistics generated in the study.**

## Reference

1 Steen VD, Medsger TA. Changes in causes of death in systemic sclerosis, 1972-2002. Ann Rheum Dis 2007;66:940–4.

2 Tamby MC, Chanseaud Y, Guillevin L, et al. New insights into the pathogenesis of systemic sclerosis. Autoimmun Rev 2003;2:152–7.

3 Valenzi E, Bulik M, Tabib T, et al. Single-cell analysis reveals fibroblast heterogeneity and myofibroblasts in systemic sclerosis-associated interstitial lung disease. Ann Rheum Dis 2019;78:1379–87.

4 Papazoglou A, Huang M, Bulik M, et al. Epigenetic Regulation of Profibrotic Macrophages in Systemic Sclerosis-Associated Interstitial Lung Disease. Arthritis Rheumatol 2022;74:2003–14.

5 Shao Y, Saredy J, Yang WY, et al. Vascular Endothelial Cells and Innate Immunity. Arterioscler Thromb Vasc Biol 2020;40:E138–52.

6 Amersfoort J, Eelen G, Carmeliet P. Immunomodulation by endothelial cells-partnering up with the immune system? Nat Rev Immunol 2022;22:576–88.

7. Lu Y, Sun Y, Xu K, et al. Editorial: Endothelial cells as innate immune cells. Front Immunol 2022;13:1–5.

8 Mendoza FA, Piera-Velazquez S, Farber JL, et al. Endothelial Cells Expressing Endothelial and Mesenchymal Cell Gene Products in Lung Tissue from Patients with Systemic Sclerosis-Associated Interstitial Lung Disease. Arthritis Rheumatol 2016;68:210–7.

9 Bian F, Lan YW, Zhao S, et al. Lung endothelial cells regulate pulmonary fibrosis through FOXF1/R-Ras signaling. Nat Commun 2023;14.

10 Granja, JM, Corces MR, Pierce SE. et al. ArchR is a scalable software package for integrative single-cell chromatin accessibility analysis. Nat Genet 2021;53:403–411.

11 Li Z, Kuppe C, Ziegler S, et al. Chromatin-accessibility estimation from single-cell ATAC-seq data with scOpen. Nat Commun 2021;12.

12 Korsunsky I, Millard N, Fan J, et al. Fast, sensitive and accurate integration of single-cell data with harmony. Nat Methods 2019;16:1289–96.

13 van Dijk D, Sharma R, Nainys J, et al. Recovering Gene Interactions from Single-Cell Data Using Data Diffusion. Cell 2018;174:716–729.e27.

14 Zhang Y, Liu T, Meyer CA, et al. Model-based analysis of ChIP-Seq (MACS). Genome Biol 2008;9.

15 Schep A, Wu B, Buenrostro J. et al. chromVAR: inferring transcription-factor-associated accessibility from single-cell epigenomic data. Nat Methods 2017;14:975–978.

16 Yu G, Wang LG, He QY. ChIP seeker: An R/Bioconductor package for ChIP peak annotation, comparison and visualization. Bioinformatics 2015;31:2382–3.

17 López-Isac E, Acosta-Herrera M, Kerick M, et al. GWAS for systemic sclerosis identifies multiple risk loci and highlights fibrotic and vasculopathy pathways. Nat Commun 2019;10:1–14.

18 Turner AW, Hu SS, Mosquera JV, et al. Single-nucleus chromatin accessibility profiling highlights regulatory mechanisms of coronary artery disease risk. Nat Genet 2022;54:804–16.

19 Purcell S, Neale B, Todd-Brown K, et al. PLINK: A tool set for whole-genome association and population-based linkage analyses. Am J Hum Genet 2007;81:559–75.

20 Neph S, Kuehn MS, Reynolds AP, et al. BEDOPS: High-performance genomic feature operations. Bioinformatics 2012;28:1919–20.

21 Quinlan AR, Hall IM. BEDTools: A flexible suite of utilities for comparing genomic features. Bioinformatics 2010;26:841–2.

22 Butler A, Hoffman P, Smibert P, et al. Integrating single-cell transcriptomic data across different conditions, technologies, and species. Nat Biotechnol 2018;36:411–20.

23 Li SF & Gong MJ. Single-cell RNA sequencing reveals the heterogeneity of endothelial cells in human dupuytren’s disease. Preprint in Researchsquare. 10.21203/rs.3.rs-3005025/v2

24 Wolock SL, Lopez R, Klein AM. Scrublet: Computational Identification of Cell Doublets in Single-Cell Transcriptomic Data. Cell Syst 2019;8:281–291.e9.

25 Domínguez Conde C, Xu C, Jarvis LB, et al. Cross-tissue immune cell analysis reveals tissue-specific features in humans. Science (80-) 2022;376.

26 Trapnell C, Cacchiarelli D, Grimsby J, Pokharel P, Li S, Morse M, et al. The dynamics and regulators of cell fate decisions are revealed by pseudotemporal ordering of single cells. Nat Biotechnol 2014. 32, 381–386.

27 La Manno G, Soldatov R, Zeisel A, et al. RNA velocity of single cells. Nature 2018;560:494–8.

28 Garcia-Alonso L, Lorenzi V, Mazzeo CI, et al. Single-cell roadmap of human gonadal development. Nature 2022;607:540–7.

29 Kuppe C, Ramirez Flores RO, Li Z, et al. Spatial multi-omic map of human myocardial infarction. Nature 2022;608:766–77.

30 Perelas A, Silver RM, Arrossi A V., et al. Systemic sclerosis-associated interstitial lung disease. Lancet Respir Med 2020;8:304–20.

31 Di Martino ML, Frau A, Losa F, et al. Role of circulating endothelial cells in assessing the severity of systemic sclerosis and predicting its clinical worsening. Sci Rep 2021;11:1–9.

32 Corces MR, Shcherbina A, Kundu S, et al. Single-cell epigenomic analyses implicate candidate causal variants at inherited risk loci for Alzheimer’s and Parkinson’s diseases. Nat Genet 2020;52:1158–68.

33 Alexi N, Inge RH, Nicole GC, et al. Brain cell type-specific enhancer-promoter interactome maps and disease-risk association. Science (80-) 2019;1139:1134–9.

34 Maurano MT, Humbert R, Rynes E, et al. Systematic localization of common disease-associated variation in regulatory DNA. Science (80-) 2012;337:1190–5.

35 Ahn Y, Sanderson BW, Klein OD, et al. Inhibition of Wnt signaling by wise (Sostdc1) and negative feedback from Shh controls tooth number and patterning. Development 2010;137:3221–31.

